# Protection following BNT162b2 booster substantially exceeds that of a fresh 2-dose vaccine: a quasi-experimental study

**DOI:** 10.1101/2021.12.19.21267933

**Authors:** Ofra Amir, Yair Goldberg, Micha Mandel, Yinon M. Bar-On, Omri Bodenheimer, Nachman Ash, Sharon Alroy-Preis, Amit Huppert, Ron Milo

## Abstract

Israel began administering a BNT162b2 booster dose to restore protection following the waning of the 2-dose vaccine. Biological studies have shown that a fresh booster leads to increased antibody levels compared to a fresh 2-dose vaccine, which may suggest increased effectiveness. To compare the real-world effectiveness of a fresh booster dose with that of a fresh 2-dose vaccine, we conduct a quasi-experimental study that compares populations that were eligible to receive the vaccine at different times due to age cutoff policies. Our analysis shows that a fresh booster increases protection against confirmed infection by 3.7 (95% CI: 2.7 to 5.2) fold compared to a fresh 2-dose vaccine.

## Main

Following observations of waning in the protection conferred by the BNT162b2 (Pfizer-BioNTech) vaccine,^1^ Israel began the administration of a third (BNT162b2 booster) dose on July 30, 2021 to individuals 60 years or older. Starting August 29, 2021, every person aged 16 or older who was vaccinated for at least 5 months was eligible to receive a booster dose. Population studies have shown that the booster is highly effective in restoring protection against infection, reducing the rate of confirmed infections and severe outcomes by several folds compared to doubly-vaccinated individuals 5 months after vaccination.^2–4^ Laboratory studies showed that a BNT162b2 booster dose significantly increased the antibody neutralization and the IgG titers levels compared to a second dose,^5,6^ which suggests possible increased protection, as increased neutralization titer could lead to increased protection against infection.^7,8^ We sought to compare the real-world effectiveness of a “fresh” BNT162b2 booster dose in preventing infection to that of a “fresh” 2-dose BNT162b2 vaccine. However, comparing the effectiveness of the booster dose with that of two “fresh” doses is challenging due to selection bias. Individuals who received the booster dose chose to vaccinate earlier, which has been shown to be correlated with high sociodemographic status.^9^

To mitigate this bias, we use a quasi-experimental study.^10^ In Israel, residents aged 16 or older were eligible to vaccinate starting February 2021, while teenagers aged 12-15 became eligible to receive the vaccine only in June 2021. We utilize the change in the age cutoff to compare the protection conferred by a fresh 2-dose vaccine to that conferred by a fresh booster dose. The quasi-experimental study is not bias-free as a controlled experiment, as prior to the vaccination campaign, the younger age group had a somewhat lower rate of confirmed infections than the 16-18 group, which is in line with prior studies showing lower infectivity and susceptibility in younger children.^11^ On the other hand, following the vaccination campaign, the older age group might have a higher indirect protection from being in an environment with higher vaccination rates. We discuss possible biases resulting from the different ages in the two cohorts in the Supplementary Appendix, as well as several sensitivity analyses exploring the magnitude of the biases. We also present a secondary analysis comparing protection of the cohorts to unvaccinated individuals in corresponding age groups.

Our main analysis compared the rates of confirmed infections between September 12, 2021 and October 9, 2021 (4th wave in Israel, which was Delta-dominant) in two cohorts: individuals aged 16-18 who received the booster dose and individuals aged 12-14 who were recently vaccinated (2 doses). Individuals in both cohorts chose to vaccinate soon after becoming eligible. For the doubly-vaccinated cohort, we only included persons who were vaccinated for less than 60 days to avoid the effect of waning immunity.^1^ We used a Poisson regression (see Supplementary Appendix) to estimate the confirmed infection rates in the two cohorts during the four-week study period. We did not analyze protection against severe disease as there were only a few cases.

The analysis shows that a fresh booster dose provides a 3.7 (95% CI: 2.7-5.2) fold increase in protection against confirmed infection compared to a fresh 2-dose vaccine (see Figure 1). The infection rate in the booster cohort was 3.3 (95% CI: 2.4-4.6) per 100,000 at-risk days, compared to 12.4 (95% CI: 11.4-14) in the doubly-vaccinated cohort.

**Figure 1:**
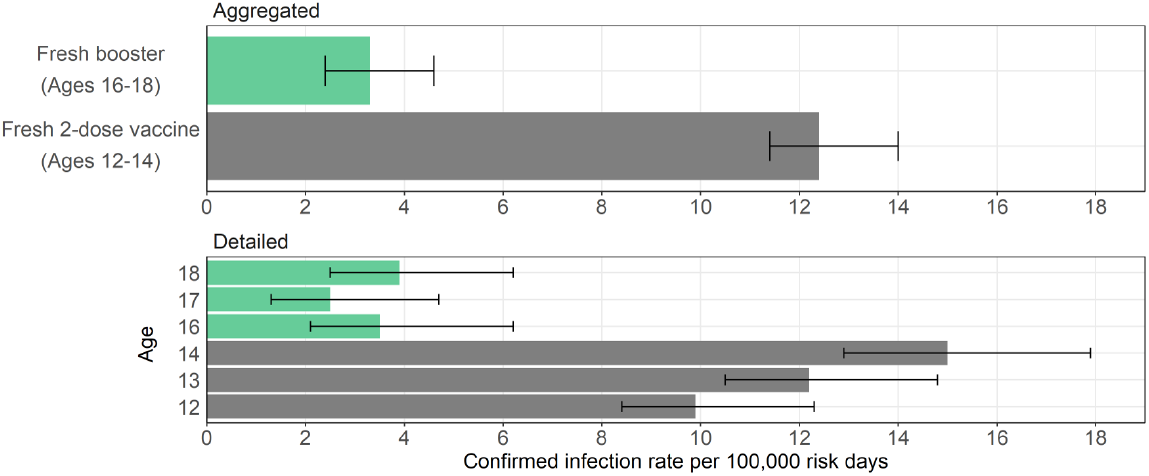
Estimated covariate-adjusted rates of confirmed infections per 100,000 at-risk days obtained from a Poisson regression analysis for the study period September 12, 2021, to October 9, 2021, stratified by cohorts. The top plot shows the results of the main analysis of the two study cohorts. The bottom plot shows covariate-adjusted rates of confirmed infections for each age group. Confidence intervals are not adjusted for multiplicity.

Our results show that a fresh booster dose of the BNT162b2 mRNA vaccine provides improved protection against confirmed infection compared to fresh two doses of the same vaccine. These results are in line with lab-based findings showing increased antibody responses both in the IgG titers as well as neutralizing antibody levels of the booster dose.^5^ Moreover, the neutralizing antibodies after the booster were superior in neutralization assay to antibodies after a second dose against both Delta and Omicron.^12–14^ Together, these findings offer a scientific basis for the inclusion of a booster dose as part of the BNT162b2 regimen.

## Data Availability

The individual-level data used in this study are sensitive and cannot be publicly shared.

## Supplementary Appendix

### Study Information

#### Ethics statement

The study was approved by the Institutional Review Board of the Sheba Medical Center. Helsinki approval number: SMC-8228-21.

#### Data sharing

The individual-level data used in this study are sensitive and cannot be publicly shared.

### Supplementary Methods

#### Description of the Data

The analysis is based on the Israel Ministry of Health’s database, as described in our previous studies.^15^ Israel has experienced four pandemic waves, with the Delta (B.1.617.2) variant being the predominant variant during the fourth wave. During the third wave, Israel initiated a very rapid vaccination campaign administering the BNT162b2 vaccine to all adult residents. The campaign was opened on December 20, 2020, initially to people aged 60 years or older, and was then gradually extended until, on February 4, 2021, all individuals aged 16 or older were eligible to receive two doses of the vaccine. After the arrival of the Delta variant to Israel, a new Covid-19 wave began in mid-June 2021. Consequently, on July 30, 2021, the administration of a third (booster) dose was approved, first for people aged 60 years or older, and later for younger age groups.^2^

Israel has a centralized health system, where each resident belongs to one of four health maintenance organizations (HMOs). Polymerase Chain Reaction (PCR) tests for SARS-CoV-2 infections as well as vaccination against the virus are provided free of charge, and are directly reported to the Ministry of Health (MoH). The MoH established a centralized Covid-19 national database containing regularly updated information on all PCR tests and results, vaccination dates, and follow-up data on all infected individuals, including the severity of disease and mortality. The MoH database also includes basic demographic information, such as sex, age, place of residency, and population sector.

#### Study Design and Population

We used a quasi-experimental design, utilizing the changes made to the vaccine eligibility age cutoffs to estimate the effectiveness of a booster dose to that of a “fresh” 2-dose vaccine. The study population included persons who were between the ages of 12-14 or 16-18 starting January 1st, had no documented positive PCR result prior to the study period, had not stayed abroad during the whole study period, and had not been vaccinated with a vaccine different from BNT162b2 before the end of the study period. We did not include the 15-year-old group since the data includes the age of individuals in one-year groups (based on their age on January 1st, 2021), and the 15-year-old group thus includes individuals who were eligible to vaccinate at different times.

In the primary analysis, the study population was divided into two cohorts. The first cohort included individuals 16-18 who recently received the booster dose, and the second cohort included individuals aged 12-14 who were doubly vaccinated for less than 60 days (to avoid waning effects). To estimate protection conferred by the 2-dose and the booster vaccines, we compared their infection rates during the four-week study period: September 12, 2021 and October 9, 2021 (4th wave in Israel, which was Delta-dominant).

In a secondary analysis, we also included a third cohort consisting of individuals aged 16-18 who were doubly vaccinated for less than 60 days. In this analysis, we used unvaccinated individuals as a reference group, and estimated the protection of each cohort compared to the corresponding unvaccinated age group during the study period.

#### Statistical Analysis

We analyzed the data using a methodology similar to that used in our previous studies.^2,3^ The number of confirmed infections and the number of days at risk during the study period were counted for each cohort. A Poisson regression model was fitted adjusting for age (by year), sex, population group (General Jewish, Arab, ultra-Orthodox Jewish), calendar week, and an exposure risk measure. The latter was calculated for each person on each follow-up day according to the proportion of new confirmed infections during the past seven days in their area of residence; this continuous measure was then divided into ten categories according to deciles (see Bar-On et al.^2^ for details). An average risk was imputed to individuals with missing data on residency.

### Secondary analysis - comparison with unvaccinated and 16-18 vaccinated

In the primary quasi-experimental analysis, biases may occur due to age differences, either because natural protection is correlated with age, or due to behavioral differences. Our secondary analysis compared the protection against confirmed infection between three cohorts: individuals aged 16-18 who received the booster dose, individuals aged 12-14 who were recently vaccinated (2 doses), and a new cohort of individuals aged 16-18 who were recently vaccinated (2 doses). The 12-14 vaccinated group is similar to the booster group in that they chose to vaccinate soon after becoming eligible. However, due to the age difference, this group also seems to be more exposed than the 16-18 group according to infections among unvaccinated individuals (see Figure S2). The 16-18 vaccinated cohort presumably has a similar exposure risk as that of the 16-18 unvaccinated cohort and does not suffer from this bias, but could introduce a behavioral bias as they were vaccinated relatively late. To measure protection, we use unvaccinated individuals as a reference group, and estimate, using a Poisson regression (see Statistical Analysis), the protection of each cohort compared to the corresponding unvaccinated age group during the four-week study period: September 12, 2021 and October 9, 2021 (4th wave in Israel, which was Delta-dominant).

The analysis shows that the booster provides a 2–3 fold increase in protection against confirmed infection compared to a fresh 2-dose vaccine (see Figure S3). Infection rates in the booster cohort were 26.3-fold (95% CI: 19.2, 36) lower than in the corresponding unvaccinated group. In the vaccinated 12-14 cohort, the increase in protection was 2.1-fold lower than in the booster cohort, with a 12.5-fold (95% CI: 11.2, 13.8) reduction in infections compared to the unvaccinated group of that age. The increase in protection in the vaccinated 16-18 cohort was 2.7-fold lower than in the booster group, with a rate of infections 9.8-fold (95% CI: 5, 16) lower than in the unvaccinated group.

### Sensitivity analyses

We conducted several sensitivity analyses. First, the 17-18 age group comprises individuals who graduated from school prior to the study period. Therefore, they may differ from the younger age groups that are exposed to other students. Indeed, we observed lower infection rates in general in the 17-18 year old unvaccinated age group. We therefore repeated the analysis removing this age group, and also compared only vaccinated 14 year old individuals with 16 year old individuals in the booster cohort, who are the closest age groups within these cohorts. These analyses show a 3.5 (95% CI: 2, 6.1) fold increase in protection for a fresh booster dose compared to a fresh 2-dose vaccine when removing the 17-18 year old age group, and a 4.6 (95% CI: 2.5, 8.1) increase when comparing only 16 year old (booster) with 14 year old (doubly-vaccinated).

Second, because booster uptake rates are higher in the General Jewish population, we conducted an additional analysis which only included this sub-population. This analysis shows a 4.1 (95% CI: 2.8, 6.2) fold improved protection for a fresh booster dose. Lastly, we analyzed confirmed infection rates over a longer study period - from September 12 to Oct 24, resulting in an estimate of a 3.5 (95% CI: 2.4, 5.2) increase in protection for the booster cohort. This analysis adds more individuals and confirmed infection events, but also includes individuals who were doubly-vaccinated later. Overall, all sensitivity analyses yielded similar results, with a fresh booster dose providing an estimated 3–5 fold increase in protection compared to fresh two doses.

### Study Limitations

The study has several limitations. We observe different confirmed infection rates for unvaccinated individuals in different age groups (see Figure S2) with higher rates up to age 16 and lower rates from age 17 and above. These differences are possibly related to the 17+ age groups graduating from school prior to the study period, which could affect their exposure and testing rates, but could also relate to different characteristics of the unvaccinated population at different ages. To account for this bias, we adjust for age by year in the regression. In a secondary analysis, we also included the vaccinated 16-18 cohort which is similar to the booster cohort with respect to age. However, this group is different in that they vaccinated later, as opposed to the booster cohort which consists of individuals who were early to vaccinate. This can reflect differences between the populations as shown in prior studies.^9^ Furthermore, prior studies have shown that younger adolescents might be less susceptible to infection than older individuals.^11^ We also observe this when examining the relevant age groups in the period of Oct-Dec 2020 (pre-Alpha dominant period in Israel), prior to their eligibility to vaccinate, where the 12–14 group had a lower confirmed infection rate than the 16–17 group by a factor of 0.8. This suggests that part of the protection in the older population in our study stems from their environment being more protected (higher vaccination rates). To address these biases we compare the booster cohort both with the vaccinated 12-14 cohort and the vaccinated 16-18 cohort, using the unvaccinated cohort in the corresponding age groups as a baseline reference. We also conducted sensitivity analyses with respect to the age groups. We note that the unvaccinated group likely includes a substantial number of undocumented recovered individuals.^16^

In addition to differences in exposure between age groups, there are also possible differences in testing rates. To account for this, we examine the number of PCR tests performed by individuals in the different cohorts (see Figure S4). We find that the vaccinated 12-14 cohort and the booster cohort have similar testing rates. Related to the secondary analysis which also included the vaccinated 16-18 cohort, we observe a somewhat lower testing rate in this group, suggesting that their protection might be overestimated. We also observe that the unvaccinated population in the 12-14 age group (used as a reference in the secondary analysis) were tested at a higher rate than the vaccinated 12-14 cohort and also at a higher rate than the unvaccinated 16-18 age group. Since the protection rates use the unvaccinated groups as reference, it is possible that the protection of the vaccinated 12-14 group is overestimated. Therefore, the increase in protection conferred by a booster dose compared to two doses might be even higher than estimated in our analysis.

Another limitation of the study is that individuals in the booster cohort chose to vaccinate early (similar to the 12-14 vaccinated cohort) and also chose to receive the booster dose, leading to a potential selection bias in the booster population. In particular, the General Jewish population had a higher uptake rate of the booster dose. Therefore, we conducted a sensitivity analysis including only this sector. We also conducted a sensitivity analysis extending the study period by two weeks, which includes a higher number of individuals who received the booster dose.

Lastly, we note that our estimates for protection reflect the real-world effectiveness of the vaccine and likely include indirect effects such as the added protection due to others being vaccinated, and not only the biological protection conferred by the vaccine. Nevertheless, all the sensitivity analyses we conducted showed that protection against confirmed infection after receiving the booster dose was substantially higher than that after two doses, suggesting that the booster dose indeed improves protection beyond that of only two doses.

---

**Figure S1.**
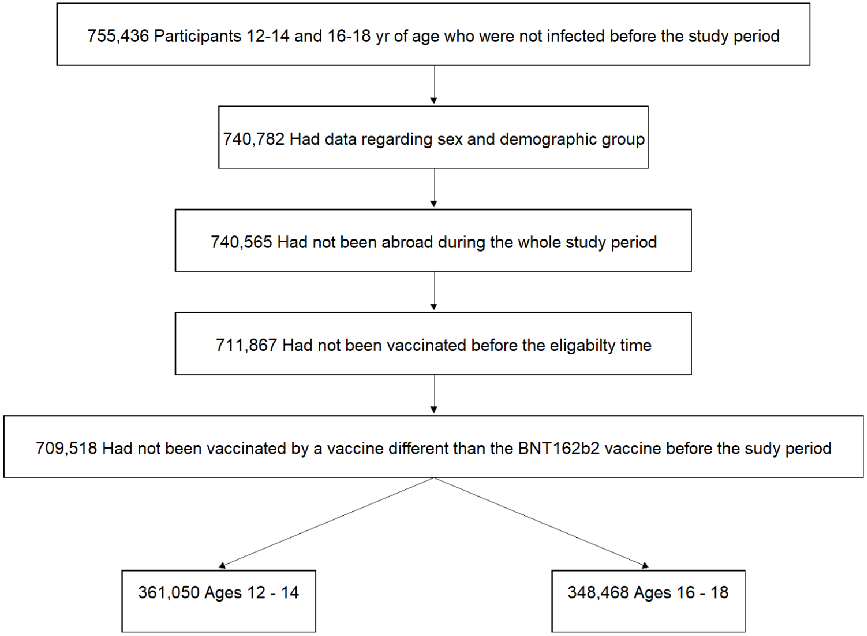
Study population. The study population included persons who were between the ages of 12-14 or 16-18, had no documented positive PCR result prior to the study period, had not stayed abroad during the whole study period, and had not been vaccinated with a vaccine different from BNT162b2 before the beginning of the study period.

**Table S1.** Demographic and clinical characteristics of the study groups. The booster group comprises individuals of ages 16-18, 14 or more days after they received the booster dose. The doubly-vaccinated groups comprise of individuals 14-60 days after they received the second dose, in ages 12-14 or 16-18. The unvaccinated groups (reference groups in the secondary analysis) comprise of individuals in ages 12-14 or 16-18 who were not vaccinated. Only person-days and events that were used in the main and secondary analyses are presented. The table presents the proportion of person-days at risk instead of the proportion of individuals. Values are presented for the study period - September 12, 2021 to October 9, 2021.

| Group | Unvaccinated 12-14<br>Person-days at risk =<br>2,837,195 |  | Unvaccinated 16-18<br>Person-days at risk =<br>1,818,163 |  | Vaccinated 12-14<br>Person-days at risk =<br>2,645,121 |  | Vaccinated 16-18<br>Person-days at risk =<br>133,613 |  | Booster 16-18<br>Person-days at risk =<br>1,716,429 |  |
| --- | --- | --- | --- | --- | --- | --- | --- | --- | --- | --- |
|  | % person<br>days at<br>risk | #<br>infections | % person<br>days at<br>risk | #<br>infections | % person<br>days at<br>risk | #<br>infections | % person<br>days at<br>risk | #<br>infections | % person<br>days at<br>risk | #<br>infections |
| Female | 51.90% | 2,805 | 53.30% | 1,038 | 51.90% | 245 | 47.40% | 7 | 49.30% | 21 |
| Male | 48.10% | 2,629 | 46.70% | 698 | 48.10% | 146 | 52.60% | 9 | 50.70% | 19 |
| General<br>Jewish | 60.50% | 2,622 | 59.90% | 930 | 76.60% | 260 | 53.30% | 11 | 88.20% | 29 |
| Arab | 15.30% | 635 | 23.00% | 303 | 7.60% | 33 | 8.90% | 0 | 5.40% | 4 |
| Ultra-Orthodo<br>x | 24.10% | 2,177 | 17.20% | 503 | 15.70% | 98 | 37.80% | 5 | 6.40% | 7 |

**Figure S2.**
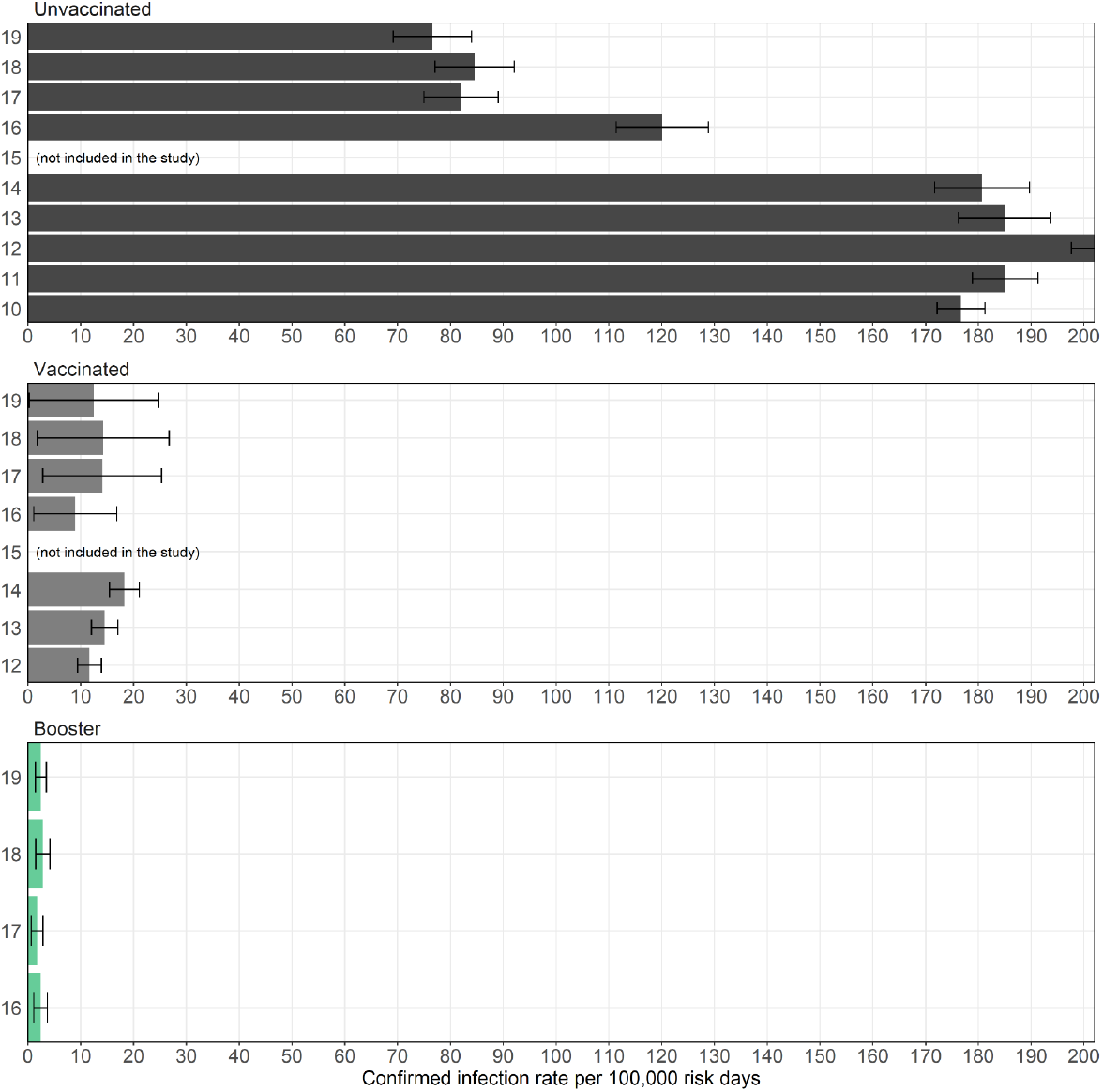
Confirmed infection rates in different age groups. Crude infection rates per 100,000 at-risk days during the study period between September 12, 2021 to October 9, 2021, stratified by cohort and age. The 15-year-old age group is not shown since it includes individuals who were eligible to vaccinate at different times.

**Figure S3.**
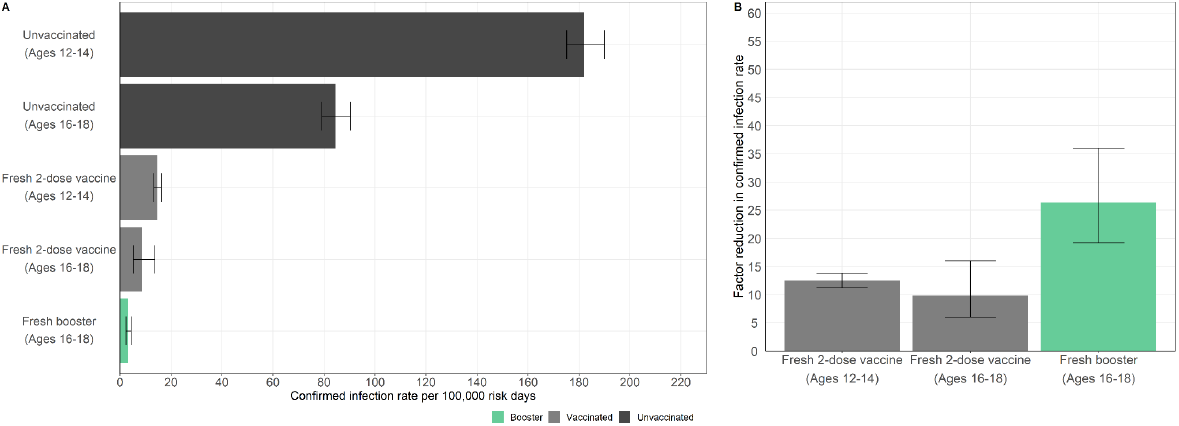
Protection compared to unvaccinated individuals. Estimated covariate-adjusted rates of confirmed infections per 100,000 at-risk days (A) and factor reductions in confirmed infections (B) obtained from a Poisson regression analysis for the study period September 12, 2021, to October 9, 2021, stratified by cohorts. Confidence intervals are not adjusted for multiplicity.

**Figure S4.**
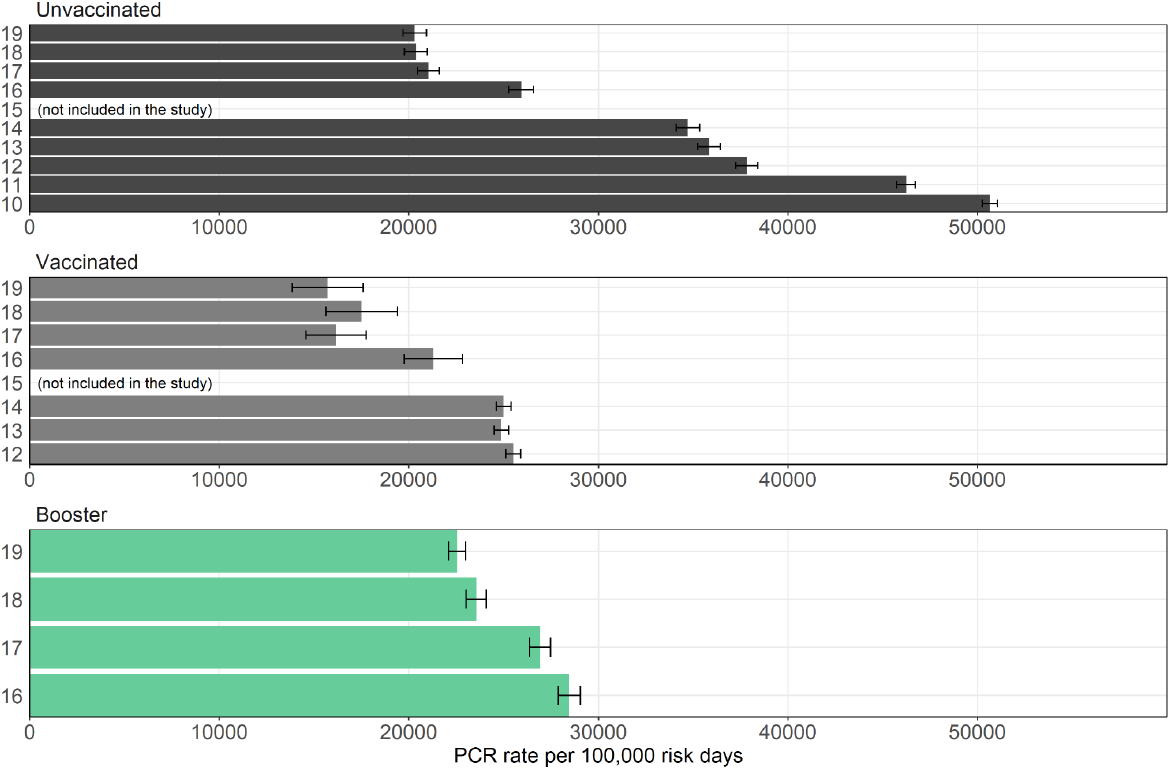
PCR testing rates in different age groups. PCR test rates per 100,000 risk days during September 2021, stratified by cohorts and ages. The 15-year-old age group is not shown since it includes individuals who were eligible to vaccinate at different times.

## References

1. Goldberg Y, Mandel M, Bar-On YM, et al. Waning Immunity after the BNT162b2 Vaccine in Israel. N Engl J Med. 2021;0(0):null. doi:10.1056/NEJMoa2114228

2. Bar-On YM, Goldberg Y, Mandel M, et al. Protection against Covid-19 by BNT162b2 Booster across Age Groups. N Engl J Med. Published online December 8, 2021:NEJMoa2115926. doi:10.1056/NEJMoa2115926

3. Bar-On YM, Goldberg Y, Mandel M, et al. Protection of BNT162b2 vaccine booster against Covid-19 in Israel. N Engl J Med. 2021;385(15):1393–1400.

4. Barda N, Dagan N, Cohen C, et al. Effectiveness of a third dose of the BNT162b2 mRNA COVID-19 vaccine for preventing severe outcomes in Israel: an observational study. The Lancet. Published online 2021.

5. Eliakim-Raz N, Leibovici-Weisman Y, Stemmer A, et al. Antibody Titers Before and After a Third Dose of the SARS-CoV-2 BNT162b2 Vaccine in Adults Aged≥ 60 Years. Jama. Published online 2021.

6. Nemet I, Kliker L, Lustig Y, et al. Third BNT162b2 Vaccination Neutralization of SARS-CoV-2 Omicron Infection.; 2021:2021.12.13.21267670. doi:10.1101/2021.12.13.21267670

7. Khoury DS, Cromer D, Reynaldi A, et al. Neutralizing antibody levels are highly predictive of immune protection from symptomatic SARS-CoV-2 infection. Nat Med. Published online 2021:1-7.

8. Cromer D, Steain M, Reynaldi A, et al. Neutralising antibody titres as predictors of protection against SARS-CoV-2 variants and the impact of boosting: a meta-analysis. Lancet Microbe. Published online 2021.

9. Rossman H, Shilo S, Meir T, Gorfine M, Shalit U, Segal E. COVID-19 dynamics after a national immunization program in Israel. Nat Med. 2021;27(6):1055–1061. doi:10.1038/s41591-021-01337-2

10. Eliopoulos GM, Harris AD, Bradham DD, et al. The Use and Interpretation of Quasi-Experimental Studies in Infectious Diseases. Clin Infect Dis. 2004;38(11):1586–1591. doi:10.1086/420936

11. Dattner I, Goldberg Y, Katriel G, et al. The role of children in the spread of COVID-19: Using household data from Bnei Brak, Israel, to estimate the relative susceptibility and infectivity of children. PLoS Comput Biol. 2021;17(2):e1008559.

12. Wilhelm A, Widera M, Grikscheit K, et al. Reduced Neutralization of SARS-CoV-2 Omicron Variant by Vaccine Sera and monoclonal antibodies. medRxiv. Published online January 1, 2021:2021.12.07.21267432. doi:10.1101/2021.12.07.21267432

13. Basile K, Rockett RJ, McPhie K, et al. Improved Neutralization of the SARS-CoV-2 Omicron Variant after Pfizer-BioNTech BNT162b2 COVID-19 Vaccine Boosting. Microbiology; 2021. doi:10.1101/2021.12.12.472252

14. Gruell H, Vanshylla K, Tober-Lau P, et al. MRNA Booster Immunization Elicits Potent Neutralizing Serum Activity against the SARS-CoV-2 Omicron Variant. Infectious Diseases (except HIV/AIDS); 2021. doi:10.1101/2021.12.14.21267769

15. Goldberg Y, Mandel M, Bar-On YM, et al. Protection and Waning of Natural and Hybrid COVID-19 Immunity. Epidemiology; 2021. doi:10.1101/2021.12.04.21267114

16. Kahn R, Schrag SJ, Verani JR, Lipsitch M. Identifying and Alleviating Bias Due to Differential Depletion of Susceptible People in Post-Marketing Evaluations of COVID-19 Vaccines. Epidemiology; 2021. doi:10.1101/2021.07.15.21260595

